# Expectancy-Value-Cost motivational theory to explore final year medical students’ research intentions and past research experience: a multicentre cross-sectional questionnaire study

**DOI:** 10.1101/2021.01.04.20244038

**Authors:** Louis Van Maele, Christelle Devos, Séverine Guisset, Sophie Leconte, Jean Macq

**Affiliations:** Institut de Recherche Santé et Société (IRSS), Université Catholique de Louvain, Brussels, Belgium; Centre Académique de Médecine Générale (CAMG), Université Catholique de Louvain, Brussels, Belgium; Institut de recherche en sciences psychologiques (IPSY), Université Catholique de Louvain, Louvain-la-Neuve, Belgium; Plateforme technologique de support en méthodologie et support statistique (SMCS), Université Catholique de Louvain, Louvain-la-Neuve, Belgium; Louvain Institute of Data Analysis and Modeling in economics and statistics (LIDAM), Université Catholique de Louvain, Louvain-la-Neuve, Belgium

## Abstract

**Objectives:** Conducting research during medical school is a commonly described way to strengthen the physician-scientists workforce. The aim of this study is to compare the strength of association of Expectancy, Value and Cost regarding a research activity with future research intentions, and to explore differences between students with or without research experience during medical school.

**Design, setting and participants:** An online questionnaire was sent to final-year medical students – who had already chosen their specialty – in three French-speaking Belgian universities with non-mandatory research programmes. Exploratory factor analysis (EFA) and multiple regression analysis were conducted.

**Main measures:** Research intention (outcome measure) was assessed using a 3-item scale. The motivational beliefs were assessed using a 10-item scale adapted from a validated scale based on the Expectancy-Value- Cost theory. Responses were recorded on a 6-point Likert scale.

**Results:** Participation rate was 28% (n=237). EFA revealed 4 factors with high internal consistency. 21.5% of students had positive research intentions (score 5 or above). Our model explained 82.8% of research intention variance (*p* < 0.001), of which three motivational beliefs had statistically significant coefficients: i) Value given to a research activity (β = 0.72, *p* < 0.001); ii) perceived Cost of a research activity (β = -0.11, *p* < 0.01); iii) Expectancy of success (β = 0.10, *p* < 0.05). Students with a positive research experience or students without research experience but who had strongly considered achieving one were 11.5 times more likely (95% CI, 5.0 – 26.2) to have positive research intentions at the end of medical school than other students.

**Conclusions:** Value given to a research activity is the key factor regarding students’ motivation to undertake research. Our study confirms the positive relationship between non-mandatory research and future research intentions, although some students without a research experience showed high motivation as well.

## INTRODUCTION

Over the past decades, academics recurrently signal the physician-scientist as an endangered species.^1-4^ Physician-scientists are defined broadly as “those with MD degrees (alone or combined with other advanced degrees) who devote a substantive percent of their professional effort to research”.^2^ They either had a certain experience in clinical practice before devoting themselves primarily to research, or are doing both at the same time, and can thus be considered to be at the intersection of care and research. This position makes them very precious, as they can help ask the right research questions or facilitate the integration of research outcomes in clinical practice.^2^ Nevertheless, reports and studies indicate a decrease of interest in research among young medical cohorts and an ageing physician-scientist workforce, which could lead to an “extinction” of the species.^3^

In order to reinforce the physician-scientist pipeline, studies suggest involving medical students in a research activity, as this is shown to increase future engagement in research.^5-8^ However, few studies have explored how and why participation in research during medical school influenced students’ future research engagement on the basis of solid theoretical foundations^.6^

Motivational theories can be of good help to understand one’s behaviour, choices, performance or volition in general and medical education.^9 10^ Ommering et al. have recently explored the effect of undergraduate medical students’ motivation for research at the start of medical school on research involvement one year later. They confirm that students act on their self-reported motivation.^11^ This suggests that fostering motivation for research could be seen as a crucial step for reinforcing the physician-scientist workforce.

Nevertheless, exploring motivation can be confusing, since definitions, operationalisation and measurement of motivation can be expressed in multiple ways.^12^ Regarding motivation for research, we have found studies using either no explicit motivation theory,^13^ social-cognitive theory,^14^ self-determination theory^11 15^ or theory of planned behaviour.^16^ Choosing the most relevant theory depends on many factors, but most importantly on the motivational outcome at stake, which Schunk et al. distinguish of four types : choice of tasks, effort, persistence and achievement^.17^

In the Belgian educational context, conducting research during or after medical school is primarily a matter of choice. One particular motivation theory relevant for exploring one’s choice of undertaking a specific task and that has not yet been used in the question of motivation for research is the Expectancy-Value-Cost theory.^18^ Initially conceived by Eccles and Wigfield,^18^ and further developed by Barron and Hulleman,^19^ they define the three major components of the theory – what they call motivational beliefs – as follows : *Expectancy of success* is defined as an individual’s belief of succeeding in a specific task in the future and is closely linked to his ability belief for that specific task. It answers the question “Can I do the task?”. *Value* is defined as a combination of an individual’s interest (or intrinsic value), perceived utility and attainment for a specific task. It answers the question “Do I want to do the task?”. *Cost* is defined as the perceived negative impact associated with performing a task, for which Barron and Hulleman propose four subcomponents: effort related to the task, effort unrelated to the task, loss of valued alternatives, and negative psychological experience. This dimension answers to the question “Am I free of barriers preventing me from investing time, energy and resources into the activity?”.

Taking all of this into account, we asked ourselves how the Expectancy-Value-Cost motivational theory can contribute to understanding final-year medical students’ research intentions, and more specifically, to what extent does Cost predict research intention compared to Expectancy and Value. Since the clinical and educational environment can already be quite overwhelming,^14^, and that there are signs of discrepancies between clinical practice and research^6^ ; since students and physician-scientists often report lack of time and perceived lower salaries as being barriers for research;^6 20^ we expect Cost to play an important role in shaping medical students’ intention for research.

Furthermore, we explored how research intentions and motivational beliefs differed when comparing students who had achieved research during medical school and those who had not. We also explored if the achievement of a research activity during medical school influenced students’ “motivational pathway” by analysing the moderation effect of a research experience on motivational beliefs. Considering the available literature on the subject, we expected students with research experience (RE) to share the same motivational pathways as students without RE, but we do expect them to have higher intentions^13 21^, higher Expectancy of success^6 14^, higher Value^22 23^, and lower Cost regarding a future research activity.

Finally, since research in our education is a non-mandatory activity, we verified if the faculties’ research opportunities met students’ demands, by exploring if students without research experience had considered doing one, and by exploring how students had perceived their research activity. As studies show comparable rates of interest and participation to research activities during medical school, we expect that faculties do meet students demands.^6^ By creating new subgroups based on these variables, we will end our analysis by exploring how research intentions and motivational beliefs differ between these different subgroups.

We expect that the answers to these questions will give researchers new ways to explore medical students’ and physicians’ research motivation and give medical schools insight on designing more evidence-based strategies to reinforce the physician-scientist pipeline.

## METHODS

### Design and participants

The study had a cross-sectional quantitative design using self-reported measures of research intentions, motivational beliefs regarding research, RE, specialty choice and socio-demographic information.

The population was composed of final-year (7^th^) medical students of all-three French-speaking medical schools of Belgium. Research is a non-mandatory academic activity that can be done in their Bachelor or Master’s program, mainly either laboratory-based (biomedical) research, or patient-based (clinical) research. In one university (ULB), research could also be achieved as a 1-month or 6- month rotation. At the time of the survey, students were on clinical rotations and had already chosen their future specialty but had not passed board exams yet.

### Conceptual framework and variables

Because “doing research” can mean a lot of things, research was defined throughout the questionnaire as “*a field of activity whose purpose is to increase knowledge in a particular field, using scientific and rigorous methods, in the laboratory or “in the field”“*. A research activity was defined as *“a research project for which you are at the origin of the scientific questioning, and in which you intervene at the different steps of the research process (from the conception of the project to the communication of the results)”*. The following framework, shown in figure 1 and based on the Expectancy-Value-Cost theory, guided our investigations.

**Figure 1.**
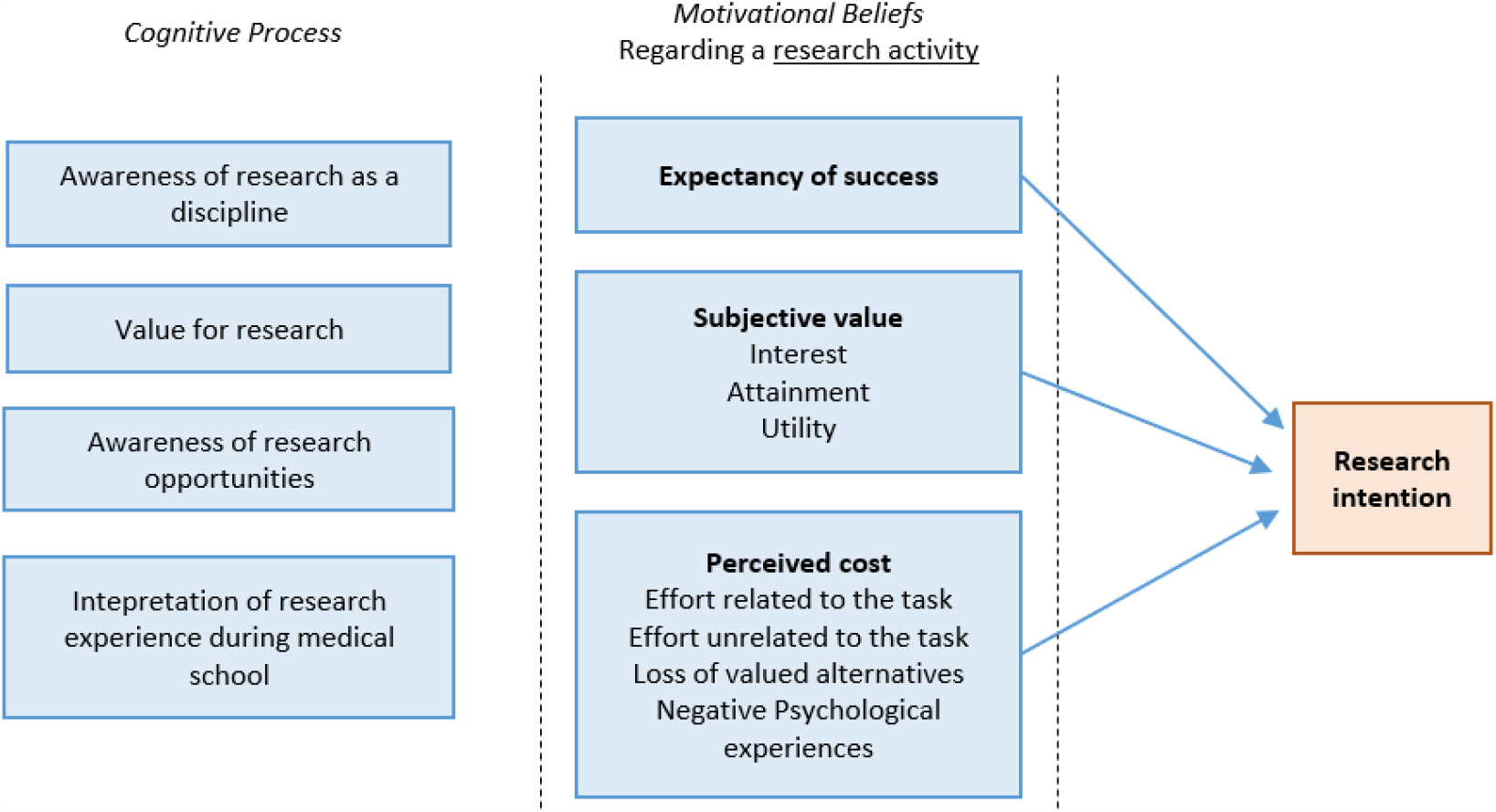
Conceptual framework.

The main outcome variable, research intention, was measured with a 3-item scale based on a theory of planned behaviour questionnaire manual.^24^ The main independent variables (Expectancy of success, Value and Cost regarding a research activity) were measured with a 10-item scale adapted from Kosovich et al.^25^ Five additional questions were added based on the conceptual framework: Awareness of research in their future specialty (1 item, self-developed) ; Value for research (how they valued research as a discipline and regarding their future clinical activities) which consisted of a 3-item scale based on the Value subscale of Kosovich et al. ; and Awareness of research opportunities (1 item, self-developed). Responses of these scales were encoded on a 6-point Likert scale going from “strongly disagree” to “strongly agree”. Four supplementary questions explored RE during medical school (2 items: yes/no and type of RE), students’ appreciation of it if they had achieved one (1 item, 5-point Likert scale), and in case they didn’t, how they had considered doing one (1 item, 5-point Likert scale).

Other variables included sociodemographic details (5 items), speciality choice (4 items), and complementary questions about future research activities and plans (11 items). All items can be found in appendix 1.

### Procedure

The 42-item questionnaire was conceived in French and pretested on five recently graduated doctors. The questionnaire was built with LimeSurvey200 and available online. In February 2017, a link to the survey was sent by email (UCLouvain and ULB) or put on the students’ virtual desktop (ULg) by local faculty administrations. Participation was anonymous and non-compulsory. Ethical approval was obtained prior to the distribution of the questionnaire. By participating to the study, students gave their informed consent to use their data.

### Analysis

We used exploratory factor analysis with oblimique rotation (direct oblimin) and Cronbach’s alpha to investigate the psychometric properties of the questionnaire. We used descriptive statistics to report age, gender, university, speciality, RE during medical school and their appreciation of it or their consideration of having done one. We calculated mean scores for each scale. We explored if scores were normally distributed by analysing skewness and kurtosis values, Shapiro-Wilk test results and histograms. We performed Spearman’s rank-order correlation and multiple regression analysis to explore the relationship between research intentions and independent variables. We explored if RE during medical school (experience over none) had moderator effects on the other variables. We finally compared differences between specific groups by calculating odds-ratio. Since the assumption of normal distribution was not met, non-parametric tests were used where indicated: Mann-Whitney-U tests when comparing two groups (groups based on RE over none), and Kruskal-Wallis tests (including post-hoc pairwise comparison test with a Bonferroni correction for multiple comparisons) when comparing six subgroups (groups based on students’ achievement, appreciation and consideration of a research activity during medical school). All data was analysed with SPSS 27.0.

### Patient and public involvement

No patients were involved.

## RESULTS

### Response rates and descriptive results

Out of 851 students invited to complete the survey, 237 completed the questionnaire for a response rate of 28%. We excluded 42 questionnaires of the analysis because responses were incomplete. A final sample of 195 questionnaires were included for analysis (23% of cohort), with specific completion rates for each university of 39% (ULB), 19% (UCLouvain), and 13% (ULg).

Descriptive results of respondents can be found in table 1.

**Table 1.** Descriptive results for all respondents and by research experience (RE)

|  |  |  |  |  |  |  |
| --- | --- | --- | --- | --- | --- | --- |
| **Variables** | **All (N = 195)** |  | **No RE (N = 125)** |  | **RE (N = 70)** |  |
|  | **N** | **%** | **N** | **%** | **N** | **%** |
| **Age (years)** | (25.0) | (1.8) | (25.2) | (2.1) | (24.8) | (1.2) |
| **Gender** |  |  |  |  |  |  |
| Female | 131 | 67% | 88 | 70% | 43 | 61% |
| Male | 64 | 33% | 37 | 30% | 27 | 39% |
| **University** |  |  |  |  |  |  |
| ULB | 92 | 47% | 62 | 50% | 30 | 43% |
| UCLouvain | 79 | 41% | 49 | 39% | 30 | 43% |
| ULg | 24 | 12% | 14 | 11% | 10 | 14% |
| **Specialty** |  |  |  |  |  |  |
| Hospital-based | 129 | 66% | 75 | 60% | 54 | 77% |
| Family medicine | 66 | 34% | 50 | 40% | 16 | 23% |
| **RE during medical school** |  |  |  |  |  |  |
| No | 125 | 64% |  |  |  |  |
| Yes | 70 | 36% |  |  |  |  |
| **Considered doing research** |  |  |  |  |  |  |
| Not at all |  |  | 49 | 39% |  |  |
| Lightly – moderately |  |  | 53 | 42% |  |  |
| (Very) strongly |  |  | 23 | 18% |  |  |
| **Feelings regarding RE** |  |  |  |  |  |  |
| (Very) negative |  |  |  |  | 7 | 10% |
| Average |  |  |  |  | 16 | 23% |
| (Very) positive |  |  |  |  | 47 | 67% |

### Scale

The exploratory factor analysis with oblique rotation (direct oblimin) was conducted on 15 items of the questionnaire. A four-factor model was retained based on our interpretation of the scree plot. This model explained 75.3% of variance with 6.5% of minimal variance explained by a factor. Details of the exploratory factor analysis can be found in appendix 2.

Table 2 shows the factor loadings after rotation. The items that group on the same factor suggest that factor 1 represents perceived Cost of a research activity (C), factor 2 represents research Awareness and value for research (A), factor 3 Expectancy of success of a research activity (E) and factor 4 Value given to a research activity (V). Cronbach’s alphas of each factor and of the intention subscale (I) are presented at the end of the same table, all being above 0.86, indicating good internal consistency.

**Table 2.** Exploratory factor analysis results and Cronbach’s alphas

|  | *Mean* | *SD* | Factors |  |  |  |  |
| --- | --- | --- | --- | --- | --- | --- | --- |
|  |  |  | 1 (C) | 2 (A) | 3 (E) | 4 (V) |  |
| Given all I will have to do in my personal or professional life, I won't have enough time to carry out a research activity | 3.6 | 1.4 | -0,96 | 0,00 | 0,15 | -0,07 |  |
| I would have to give up too many things if I wanted to achieve a research activity | 3.6 | 1.3 | -0,84 | 0,02 | -0,02 | -0,05 |  |
| I would feel incapable of devoting the necessary amount of time to achieve a research activity | 3.3 | 1.3 | -0,73 | -0,06 | -0,04 | 0,10 |  |
| Conducting a research activity would require too much time | 4.2 | 1.2 | -0,66 | 0,05 | -0,07 | -0,02 |  |
| TOTAL COST | 3.7 | 1.1 |  |  |  |  |  |
| I think research performed within the field of my future specialty will be useful for my clinical activity | 5.2 | 1.0 | -0,06 | 0,77 | -0,08 | 0,15 |  |
| It is important for me that research is done in the field of my future specialty | 5.1 | 1.2 | -0,04 | 0,76 | -0,04 | 0,07 |  |
| I think research is achieved in the field of my future specialty | 5.2 | 1.1 | 0,06 | 0,75 | 0,07 | -0,19 |  |
| I show interest towards research within the field of my future specialty | 5.0 | 1.1 | 0,06 | 0,69 | 0,03 | 0,16 |  |
| I think there are opportunities to conduct research in the field of my future specialty | 4.8 | 1.2 | 0,14 | 0,56 | 0,29 | -0,21 |  |
| TOTAL AWARENESS | 5.1 | 0.9 |  |  |  |  |  |
| I believe I could successfully carry out a research activity | 4.1 | 1.3 | 0,04 | -0,11 | 0,89 | 0,08 |  |
| I feel capable of achieving a research activity | 4.2 | 1.4 | 0,11 | 0,03 | 0,79 | 0,09 |  |
| I am certain I could understand how to conduct a research activity | 4.0 | 1.3 | -0,06 | 0,07 | 0,67 | -0,01 |  |
| TOTAL EXPECTANCY OF SUCCESS | 4.1 | 1.2 |  |  |  |  |  |
| Performing a research activity would be important for me | 3.8 | 1.4 | 0,19 | 0,15 | 0,23 | 0,66 |  |
| I would be interested in performing a research activity | 4.2 | 1.6 | 0,27 | 0,15 | 0,27 | 0,50 |  |
| I think conducting a research activity would be useful for my activity/project/professional goal | 4.3 | 1.3 | 0,11 | 0,18 | 0,24 | 0,48 |  |
| TOTAL VALUE | 4.1 | 1.3 |  |  |  |  |  |
| Eigenvalue (initial) |  |  | 7.22 | 2.02 | 1.09 | 0.97 |  |
| % variance explained (initial) |  |  | 48.1% | 13.5% | 7.3% | 6.5% |  |
| I want to perform a research activity in the future | 4.0 | 1.6 |  |  |  |  |  |
| I intend to perform a research activity in the future | 3.7 | 1.5 |  |  |  |  |  |
| I have planned to achieve a research activity in the future | 3.2 | 1.4 |  |  |  |  |  |
| TOTAL RESEARCH INTENTION | 3.6 | 1.4 |  |  |  |  |  |
|  |  |  | I | C | A | E | V |
| Number of items |  |  | 3 | 4 | 5 | 3 | 3 |
| Cronbach's alpha |  |  | 0.94 | 0.88 | 0.86 | 0.86 | 0.90 |

NB : Items were created in French and translated in English for the purpose of publishing

### Research intentions and motivational beliefs

The mean score of students’ research intention and motivational beliefs can be found in table 2. The mean score of intention was medium (3.6). Quartiles were calculated at the following scores: 2.7, 4.0 and 4.7. 21.5% of students scored at 5 or above (agree or strongly agree with the intention to do research). Twenty-eight students (14.4% of total) reported having already planned research for their future. Mean scores on motivational beliefs and awareness were higher (ranging from 3.7 to 5.1). All variables had statistically significant correlations between each other, as shown by the results of Spearman’s rank-order correlations in appendix 3.

We ran a hierarchical multiple regression to determine if the addition of research experience during medical school and motivational beliefs improved the prediction of research intention over and above age, gender, university and specialty choice alone. The full model including all variables was predictive of research intention (model 7, with details in table 3), predicting 82.8% of research intention variance (as represented by R^2^), R^2^ = 0.828, F(10, 184) = 88.883, p < 0.0005; adjusted R^2^ = 0.819. Assumptions for multiple regression analysis were verified. The addition of motivational factors led, altogether, to a statistically significant increase in explained variance of 57.5%. Details of each regression model can be found in appendix 4.

**Table 3.** Multiple regression results (full model). Research intention as dependent variable

|  |  |  |  |  |  |  |
| --- | --- | --- | --- | --- | --- | --- |
| **Variables** | **B** | **SE B** | **Beta** | **t** | **P values** | **95% CI for B** |
| (Intercept) | -0,71 | 0,71 |  | -1,00 | 0,316 | (-2.12 ; 0.69) |
| **Age** (years) | 0,04 | 0,02 | 0,05 | 1,58 | 0,115 | (-0.01 ; 0.09) |
| **Gender** |  |  |  |  |  |  |
| Female |  |  |  |  |  |  |
| Male | 0,08 | 0,10 | 0,03 | 0,79 | 0,432 | (-0.12 ; 0.27) |
| **University** |  |  |  |  |  |  |
| ULB |  |  |  |  |  |  |
| UCLouvain | -0,11 | 0,09 | -0,04 | -1,14 | 0,256 | (-0.29 ; 0.08) |
| ULg | 0,10 | 0,14 | 0,02 | 0,71 | 0,476 | (-0.18 ; 0.38) |
| **Specialty** |  |  |  |  |  |  |
| Hospital-based |  |  |  |  |  |  |
| Family Medicine | -0,08 | 0,10 | -0,03 | -0,76 | 0,451 | (-0.28 ; 0.12) |
| **RE during medical school** |  |  |  |  |  |  |
| No |  |  |  |  |  |  |
| Yes | 0,16 | 0,10 | 0,05 | 1,60 | 0,112 | (-0.04 ; 0.35) |
| **Research Awareness** | 0,02 | 0,06 | 0,02 | 0,40 | 0,687 | (-0.10 ; 0.15) |
| **Expectancy of success\*** | **0,12** | 0,05 | **0,10** | 2,14 | **0,034** | (0.01 ; 0.23) |
| **Value\*\*** | **0,77** | 0,05 | **0,72** | 14,82 | **< 0,001** | (0.67 ; 0.87) |
| **Cost\*** | **-0,14** | 0,05 | **-0,11** | -2,69 | **0,008** | (-0.24 ; -0.04) |

RE = Research experience ; Beta : standardized coefficients
\*p < 0.05 ; \*\* p < 0.001

RE during medical school was a statistically significant predictor of research intention up until the last model, which added the variable “Value” to the model. This could be explained by a problem with multicollinearity. There were no statistically significant moderator effects of RE during medical school (experience over none) on the other variables.

These results show that all-three motivational beliefs had statistically significant coefficients in the model predicting research intention scores when controlling for profile variables, specialty choice, past experience and research awareness, but that the strength of each relationship varies. Value has the strongest relationship with research intention as the Beta shows that a change of +1 standard deviation in the Value score will result in a change of +0.72 standard deviation in the intention score, all else being equal. The same reasoning applied to Expectancy of success will result in a change of +0.10 SD in the intention score, as for Cost, it would result in a change of -0.11 SD in the intention score.

### Research during medical school

Seventy respondents (36% of respondents; 33% of ULB respondents, 38% of UCLouvain, 42% of ULg) declared having done a research activity during medical school, while faculty administrations reported that 164 out of the cohort of 851 students (19% of total students ; 21% of ULB students, 24% of UCLouvain, 8% of ULg) had achieved one.

Students who had a research experience during medical school were 5.3 times more likely (95% CI, 2.5 – 11.0) to have positive research intentions at the end of medical school (score 5 or above) than students without research experience. Mann-Whitney-U tests revealed that these students had higher scores for Expectancy and Value and lower for Cost compared to students who hadn’t done research (2-sided tests with significance level of 0.05 adjusted by the Bonferroni correction for multiple tests).

We further divided these two groups based on the following. For students without a RE, the division was based on how they considered doing research during medical school (not at all, lightly or moderately, strongly or very strongly). For students with a RE, the division was based on the appreciation they had regarding their RE (very negative or negative, average, positive or very positive). Scores for each of these six subgroups can be found in table 4. Comparing the scores of these 6 subgroups, Kruskal-Wallis tests revealed statistically significant differences in distribution of scores across all 6 subgroups and for every variable. Post-hoc pairwise comparison tests revealed no statistically significant differences in distributions of scores between students who had a positive RE and students who hadn’t done research but who had strongly or very strongly considered it at some point, be it for the intention score or any motivational belief score. These two subgroups did have statistically significant differences in distributions of scores with the other four subgroups, for research intention and value. Figure 2 shows these results for research intention.

**Table 4.**
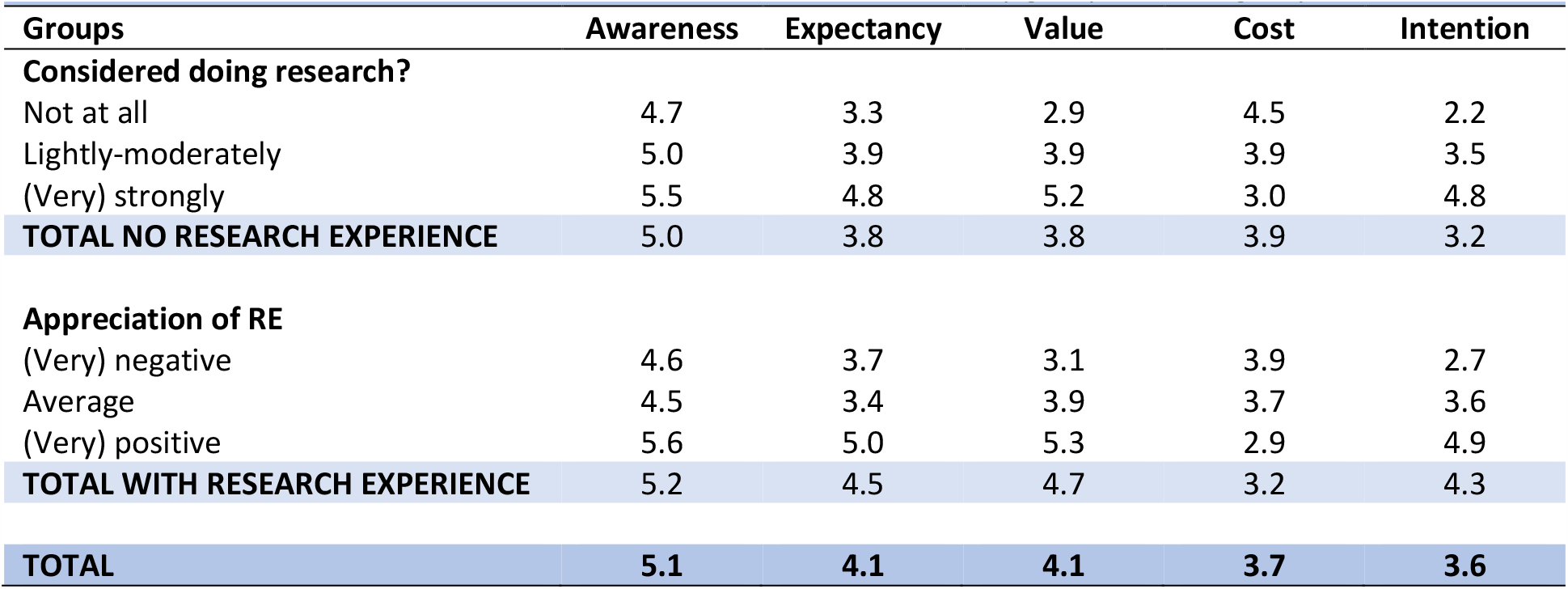
Mean scores for research intention and motivational beliefs by groups and subgroups

|  |  |  |  |  |  |
| --- | --- | --- | --- | --- | --- |
| **Groups** | **Awareness** | **Expectancy** | **Value** | **Cost** | **Intention** |
| **Considered doing research?** |  |  |  |  |  |
| Not at all | 4.7 | 3.3 | 2.9 | 4.5 | 2.2 |
| Lightly-moderately | 5.0 | 3.9 | 3.9 | 3.9 | 3.5 |
| (Very) strongly | 5.5 | 4.8 | 5.2 | 3.0 | 4.8 |
| **TOTAL NO RESEARCH EXPERIENCE** | **5.0** | **3.8** | **3.8** | **3.9** | **3.2** |
| **Appreciation of RE** |  |  |  |  |  |
| (Very) negative | 4.6 | 3.7 | 3.1 | 3.9 | 2.7 |
| Average | 4.5 | 3.4 | 3.9 | 3.7 | 3.6 |
| (Very) positive | 5.6 | 5.0 | 5.3 | 2.9 | 4.9 |
| **TOTAL WITH RESEARCH EXPERIENCE** | **5.2** | **4.5** | **4.7** | **3.2** | **4.3** |
| **TOTAL** | **5.1** | **4.1** | **4.1** | **3.7** | **3.6** |

**Figure 2.**
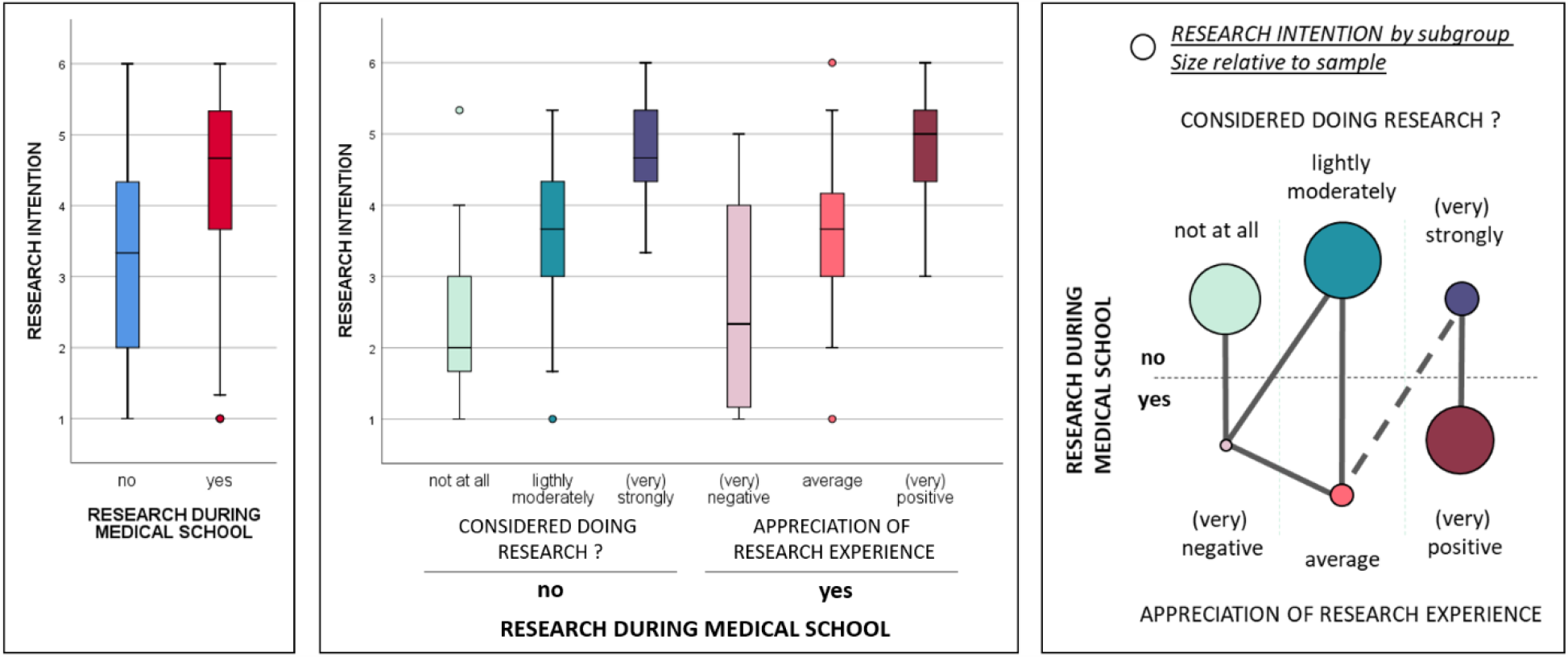
Research intention scores distributions by subgroups based on research experience during medical school, appreciation of past research experience and past research consideration. On the left, the graph shows research intentions scores with box-plots for the two main groups. In the middle, the graph shows research intentions scores with box-plots for each subgroup. On the right, the figure shows Kruskal-Wallis tests results regarding research intention. Each node represents a specific subgroup (size of the node relative to sample size). Groups that are not linked had statistically significant differences in distribution of scores (2-sided tests with significance level of 0.05 adjusted by the Bonferroni correction for multiple tests). Full lines link groups without any statistically significant differences in distribution of scores. Dashed lines link groups with statistically significant differences in distributions on a single test, but statistically insignificant after Bonferroni correction.

Only a minor number of students without RE had strongly considered doing one, and a majority of students with a RE had a positive appreciation of it. Students with a positive RE or without RE but who had strongly considered it were 11.5 times more likely (95% CI, 5.0 – 26.2) to have positive research intentions at the end of medical school (score 5 or above) than other students.

## DISCUSSION

### Principal findings

This study assessed research intention of final year medical students in all three French-speaking medical faculties of Belgium and explored how motivation differed between students who had achieved a research activity during medical school and those who had not. The originality of this study was to use the Expectancy-Value-Cost theory to assess factors associated with research intention. Our results showed that our questionnaire was relevant for investigating research intention, our model predicting 82.8% of research intention variance (*p* < 0.001). Cost had only a minor, but statistically significant, negative predictive effect on research intention score – as opposed to Value which had the highest coefficient of our model. Students had a medium mean intention score (3.6 on a 6-point Likert scale) with 21.5% of students expressing a positive research intention (score 5 or above). Students who had a RE during medical school were 5.3 times more likely (95% CI, 2.5 – 11.0) to have positive research intentions at the end of medical school (score 5 or above). They shared the same “motivational pathway” as students without RE, hence having more favourable scores in motivational beliefs as their intention score was higher. However, this dichotomisation between students with or without RE may be reductive as our results show that similarities between subgroups exist when considering how much students without RE had considered achieving one or not, and how much students with RE had a positive or negative appreciation of it. Students with a positive RE or without RE but who had strongly considered it were 11.5 times more likely (95% CI, 5.0 – 26.2) to have positive research intentions at the end of medical school (score 5 or above) than other students.

### Expectancy-Value-Cost theory and research intention

Keeping in mind that research intention is merely a reflection of actual future engagement in research, our findings confirm that the type of motivation matters.^11^ The small but statistically significant contribution of Expectancy of success is not surprising since it is described as being more related to performance outcomes.^19^ Cost however has been described as predicting both types of outcomes (performance and choice),^19^ but has a comparable – although negative – impact on research intention than Expectancy of success. We expected Cost to be an important motivational belief against a research activity considering the amount of time one should invest in research and the efforts students already have to make for learning and practising medicine. However, besides the fact that medical students may not be the average university student, it is possible that final year medical students underestimate Cost, as they may not appreciate the true workload of clinical or high-level research activities yet. Another explanation when compared with US medical students, for whom educational debt is often cited as a barrier for research-oriented careers since students perceive them as less lucrative^16^, is the near-absence of educational indebtedness in our setting since access to medical school and university in Belgium is fairly affordable.

That said, our results show that Value for a research activity is the key motivational belief regarding research intention in final year medical students. In the Expectancy-Value-Cost theory, Value for an activity is a combination of how a person believes achieving this activity is important for himself, in relation to valued aspects of his identity (attainment), is useful for reaching specific short- or long-term goals (utility) or is inherently joyful for its own sake (interest). Although the utility subcomponent is often cited as the main reason for medical students to engage in research (in order to secure a competitive residency spot or to increase access to an academic career)^6^, Ommering et al. have argued that intrinsic motivation (interest and enjoyment) influences medical student research involvement over and beyond extrinsic motivation (such as utility).^11 26^ The same authors have found that intrinsic motivation was influenced by self-efficacy beliefs, perceptions of research, curiosity and the need for challenge.^27^ Our findings also show that Value is associated with research awareness and Expectancy of success, but is also associated with Cost. Considering all this, we can justify focusing on Value and intrinsic motivation in order to increase research involvement. However, we have to keep in mind that all motivational beliefs are interconnected, and that some that might not be important for the decision of getting involved in research might well be for his performance or persistence in achieving a research activity.

### Research experience during medical school

When comparing our results to Amgad et al.’s systematic review and meta-analysis, we find that our population has a lower rate of research experience and a lower intention to undertake research in the future.^6^ Explaining these differences is not an easy task, as comparing research intentions and experiences can be difficult due to selected activities, thresholds and choice of wording of survey questions. Nevertheless, it is possible that French-speaking medical schools in Belgium have less of a research culture among their programs than what can be found in other countries, especially English-speaking ones.

Our results also show an increased likelihood of reporting positive research intentions after achieving research during medical school, compared to that same study.^6^ Considering the fact that reverse causality cannot be excluded due to our research design, one explanation for this would be that a research activity strengthens pre-existing research intentions, and does so even more in a low-level research culture.^6 11 28^ Besides, this effect has been cited as being even more plausible in the case of voluntary research, although elective research during medical school has been argued as being beneficial over compulsory research in terms of motivation and research outputs.^6 7^ Going beyond the elective-mandatory debate, our results show that motivational beliefs and future research intention differ if the student perceived his research experience as being negative, average or positive, thus leading to question ourselves on the conditions of such appreciation.

Finally, we show that offer seems to meet demand. We can say this by comparing the size of the different subgroups based on their consideration and appreciation of a research activity. A vast majority of students without a RE did not consider doing one, and a majority of students with RE had a positive appreciation of it. That said, dichotomising students on the basis of their research achievement may be reductive. Students without a research experience but who had considered doing one at some point showed nearly as much research intention than students with a positive research experience. We could hypothesise that for these students that had considered doing research, their pre-existing research intentions remain intact through their educational journey to graduation. Different reasons could explain why these students didn’t engage in a research activity such as lack of opportunities or competing interests, but the real question might not be there. As the achievement of a research activity might not seem as essential as it looks in order to increase research intention at the end of medical school, one could simply ask: would make students consider doing a research activity during medical school be enough? The answer to this question would need more research aimed at comparing these two subgroups on actual research involvement. But making students consider doing research is certainly a first step in promoting research, our results putting forward some arguments for that strategy.

### Limitations

Our study has some limitations and strengths. The cross-sectional design implies that no causal relation can be inferred by our data. It is nevertheless a theory-driven investigation, integrating how one perceives a research activity as costly, which has not yet been compared to other motivational factors in previous studies. Our sample had a higher ratio of student having achieved research during medical school compared with official faculty numbers, which suggests a potential participation bias, particularly in the university with the least participation rate (ULg). However, our sample showed an important proportion of students with low research intentions, suggesting we had managed to gather answers from a sufficiently diverse set of student profiles. Finally, our study was multicentric, thus going beyond local specific institutional research cultures or research programmes, but these differences went only up to a certain point as we stayed in the French-speaking institutions and that access to medical school is less selective and costly than in other contexts.

### Implications and further research

Our results point out the importance of how medical students value a research activity, and highlight the fact that achieving a research activity does not say all, as some students without that type of experience show high levels of motivation. Exploring students’ appreciation of a research experience and how much they considered doing research is a simple way to assess if offer meets demand in the case of a program with non-mandatory research activities. But promoting research is not about finding that balance, the goal being to attract more and more students to research. Considering the key role of Value, we invite medical educators and researchers to reflect on how research is introduced to students and how they benefit from adequate assistance and guidance in discovering what a research activity consists of. We believe this questions our relationship to knowledge and the similarities between the researcher and the clinician’s identity and purpose. Triggering reflexivity of medical students on epistemology and opening their eyes on all what research in medicine is – clinical, epidemiological, but also social, psychological, and so on – could positively influence students’ Value for a research activity.

New questions arise from this study that could benefit further research. First, linking motivational beliefs, intention for research and actual research involvement by causal relations seems to be an important step to bring more evidence on the usefulness of this motivational theory. This would imply prospective study designs. Second, exploring how motivational beliefs and intentions evolve over a student’s educational journey would help to understand how best to motivate students at each specific stage. We make the hypothesis that Value and Cost are two beliefs that can change substantially across the professional identity formation of both the doctor and the scientist. Finally, in order to understand more precisely why medical students consider doing research in the first place and what makes research during medical school a positive experience, studies using qualitative methods could help institutions optimise their research programs in order to maximise the benefits for their students.

## Conclusion

The Expectancy-Value-Cost theory has been shown to be particularly relevant for studying final year medical students’ research intention. Value for a research activity represents a key motivational belief in this matter, which justifies that efforts of educators and researchers are directed towards that component in order to increase research engagement. Students having achieved a research activity during medical school express more favourable motivational beliefs than students who have not, provided that they perceive this activity as being a positive experience. One exception: students without a research experience but who had strongly considered doing one express equivalent research intentions. Making students simply consider doing research in the first place seems to be an efficient first step in order to reinforce the physician-scientist workforce.

## Supporting information

STROBE Checklist

## Data Availability

Data are available in a public, open access repository (UCLouvain Dataverse)

https://doi.org/10.14428/DVN/FT2TX8

## AUTHOR STATEMENTS

### Ethical approval

This study was approved by the ethical review board of the Université Catholique de Louvain (Centre d’Ethique hospitalo-facultaire) : reference 2017/26 JAN/045.

### Data statement

Data are available in a public, open access repository (UCLouvain Dataverse) :

DOI : https://doi.org/10.14428/DVN/FT2TX8

### Author contributions

LVM, SL, CD and JM contributed to the design of the study. LVM performed the data analysis. SG supervised data analysis. LVM, CD, SG and JM interpreted the data. LVM wrote the manuscript. All authors contributed to the revision of the manuscript and approved the final version for publication.

## Acknowledgements

The authors wish to thank Coralie Theys for her critical discussions during the research process, as well as Stéphanie Blockx and local faculty administrations for facilitating administrative procedures.

