## appendix 1 for "Expectancy-Value-Cost motivational theory to explore final year medical students’ research intentions and past research experience: a multicentre cross-sectional questionnaire study"

**APPENDIX 1 – QUESTIONNAIRE**

**French (original) version**

| ID | ORDRE (#) | QUESTIONS | REPONSES |
|---|---|---|---|
| **SD1** | 1 | Quel est votre âge ? | |
| **SD2** | 2 | Quel est votre genre ? | M ; F |
| **SD3** | 3 | À quelle université êtes-vous actuellement inscrit ? | UCL ; ULB ; ULg |
| **SD4** | 4 | Quel est le niveau de diplôme le plus élevé obtenu par votre mère ? | Primaire ; Secondaire inférieur ; Secondaire supérieur ; Supérieur $1^{er}$ cycle (bachelier universitaire, haute école) ; Supérieur $2^{e}$ cycle (master) ; Supérieur $3^{e}$ cycle (doctorat) ; autre : |
| **SD5** | 5 | Quel est le niveau de diplôme le plus élevé obtenu par votre père ? | Idem Q4 |
| | | **CHOIX DE SPÉCIALITÉ** | |
| **SC1** | 6 | Quel est votre choix de spécialité ? | anatomie pathologique ; anesthésie-réanimation ; biologie clinique ; chirurgie générale ; chirurgie – orthopédie ; chirurgie – urologie ; chirurgie – plastique ; chirurgie – neurochirurgie ; dermatologie ; gynécologie-obstétrique ; médecine interne (sauf gériatrie et oncologie) ; médecine interne – gériatrie ; médecine interne – oncologie ; médecine générale ; médecine légale ; médecine nucléaire ; médecine physique ; neurologie ; ophtalmologie ; ORL ; pédiatrie ; pédopsychiatrie ; psychiatrie ; radiologie ; radiothérapie ; stomatologie ; urgences ; autre : |
| **(SC2)** | 7 | Avez-vous hésité avec la médecine générale ? | 1= Pas du tout ; 2= légèrement ; 3= moyennement ; 4= fortement ; 5= très fortement |
| **SC3** | 8 | Avez-vous hésité avec une autre spécialité (hors médecine générale) ? | 1= Pas du tout ; 2= légèrement ; 3= moyennement ; 4= fortement ; 5= très fortement |
| **(SC4)** | 9 | Avec quelle spécialité avez-vous le plus hésité ? | Idem Q6 (sauf MG) |
| | | **PARCOURS ACADÉMIQUE** | |

| | | | |
|---|---|---|---|
| **RE1** | 10 | Avez-vous un moment envisagé de réaliser une activité de recherche durant vos études de médecine (type mémoire, étudiant chercheur, etc.) ? | 1= Pas du tout ; 2= légèrement ; 3= moyennement ; 4= fortement ; 5= très fortement |
| **RE2** | 11 | Avez-vous réalisé une activité de recherche durant vos études de médecine (type mémoire, étudiant chercheur, etc.) ? | Oui ; non |
| **(RE3)** | 12 | De quel type d'activité s'agissait-il ? | Mémoire de recherche clinique ; étudiant chercheur ; autre : |
| **(RE4)** | 13 | Quel est votre ressenti général sur cette expérience ? | 1= Très négatif ; 2= négatif ; 3= moyen ; 4= positif ; 5= très positif |
| | | **PERCEPTION ENVERS LA RECHERCHE**<br>Cette section porte sur la façon dont vous percevez la recherche qui est réalisée dans votre future spécialité en général.<br>Par recherche, nous entendons un domaine d'activité ayant pour finalité l'accroissement des connaissances dans un domaine particulier, au moyen de méthodes scientifiques et rigoureuses, en laboratoire ou « sur le terrain ». | |
| | | Indiquez votre degré d'appréciation de chaque élément ci-dessous : | |
| | | **Reconnaissance (Awareness)** | |
| **R-A1** | 14 | Je pense que de la recherche est réalisée dans le domaine de ma future spécialité | 1= Pas du tout d'accord ; 2= pas d'accord ; 3= plutôt pas d'accord ; 4= plutôt d'accord ; 5= d'accord ; 6= tout à fait d'accord |
| | | **Valeur (Value)** | |
| **R-V1** | 15 | Il est important pour moi que de la recherche soit réalisée dans le domaine de ma future spécialité | Idem Q14 |
| **R-V2** | 16 | Je porte de l'intérêt envers la recherche réalisée dans le domaine de ma future spécialité | Idem Q14 |
| **R-V3** | 17 | Je pense que la recherche réalisée dans le domaine de ma future spécialité est utile pour ma future activité clinique | Idem Q14 |

| | | | |
|---|---|---|---|
| | | **PERCEPTION ENVERS UNE ACTIVITÉ DE RECHERCHE**<br>Cette section porte sur la façon dont vous percevez une activité de recherche que vous pourriez potentiellement réaliser dans le futur.<br>Par activité de recherche, nous entendons un projet de recherche pour lequel vous vous situez à l'origine du questionnement scientifique, et dans lequel vous intervenez aux différents stades du processus de recherche (de la conception du projet à la communication des résultats) | |
| | | Indiquez votre degré d'appréciation de chaque élément ci-dessous : | |
| | | **Reconnaissance (Awareness)** | |
| **RA-A1** | 18 | Je pense qu'il existe des opportunités pour réaliser une activité de recherche dans ma future spécialité | Idem Q14 |
| | | **Attente de réussite (Expectancy)** | |
| **RA-E1** | 19 | Je me sens capable de réaliser une activité de recherche | Idem Q14 |
| **RA-E2** | 24 | Je crois pouvoir réaliser une activité de recherche avec succès | Idem Q14 |
| **RA-E3** | 27 | Je suis certain de pouvoir comprendre la manière de réaliser une activité de recherche | Idem Q14 |
| | | **Valeur (Value)** | |
| **RA-V1** | 22 | Réaliser une activité de recherche serait important pour moi | Idem Q14 |
| **RA-V2** | 20 | Cela m'intéresserait de réaliser une activité de recherche | Idem Q14 |
| **RA-V3** | 26 | Je pense que réaliser une activité de recherche serait utile pour mon activité/projet/but professionnel | Idem Q14 |
| | | **Cout (Cost)** | |
| **RA-C1** | 21 | Réaliser une activité de recherche demanderait trop de temps | Idem Q14 |

| | | | |
|---|---|---|---|
| **RA-C2** | 28 | Vu tout ce que je devrai faire dans mon futur personnel ou professionnel, je n'aurai pas assez de temps pour réaliser une activité de recherche | Idem Q14 |
| **RA-C3** | 23 | Je me sentirais incapable de consacrer le temps nécessaire à la réalisation d'une activité de recherche | Idem Q14 |
| **RA-C4** | 25 | Je devrais renoncer à trop de choses si je voulais réaliser une activité de recherche | Idem Q14 |
| | | **INTENTION ENVERS UNE ACTIVITÉ DE RECHERCHE** | |
| | | Indiquez votre degré d'appréciation de chaque élément ci-dessous : | |
| **RA-I1** | 29 | J'ai envie de réaliser une activité de recherche dans le futur | Idem Q14 |
| **RA-I2** | 30 | J'ai l'intention de réaliser une activité de recherche dans le futur | Idem Q14 |
| **RA-I3** | 31 | J'ai prévu de réaliser une activité de recherche dans le futur | Idem Q14 |
| **RA-O1** | 32 | Comment estimez-vous vos chances de réaliser une activité de recherche dans le futur ? | 1= extrêmement faibles ; 2= très faibles ; 3= assez faibles ; 4= moyenne ; 5= assez fortes ; 6= très fortes ; 7= extrêmement fortes |
| **RA-O2** | 33 | Est-il déjà prévu que vous réalisiez une activité de recherche dans le futur ? | Oui ; non ; autre |
| **RA-O3** | 34 | A quel moment préféreriez-vous réaliser une activité de recherche ? | pendant l'assistanat ; en début de carrière médicale ; en cours de carrière médicale ; en fin de carrière médicale ; autre : |
| **RA-O4** | 35 | A quel degré de temps de travail préféreriez-vous réaliser une activité de recherche ? | <1/10 ; 1/10 ; 2/10 ; 3/10 ; 4/10 ; 5/10 ; 6/10 ; 7/10 ; 8/10 ; 9/10 ; 10/10 ; autre : |
| **RA-O5** | 36 | De quelle manière préfériez-vous réaliser une activité de recherche ? | activité professionnelle unique ; activité professionnelle principale, avec une pratique clinique comme activité secondaire ; activité professionnelle secondaire, avec une pratique clinique comme activité principale ; autre : |

| | | | |
|---|---|---|---|
| **RA-O6** | 37 | Dans quel type d’environnement de travail préféreriez-vous réaliser une activité de recherche ? | Université ; société savante/autre regroupement scientifique professionnel ; société privée (ex. : pharmaceutique, autre) ; pratique/hopital/réseau local ; autre : |
| **RA-O7** | 38 | Quel type d'activité de recherche préféreriez-vous réaliser ? | Travail de fin d’étude orienté recherche ; Thèse de doctorat ; Activité professionnelle (unique ou complémentaire) ; Autre : |
| | | **RAISON** | |
| **RA-O8** | 39 | Quelle serait votre raison principale à entamer une activité de recherche ? | -argent : je pense que faire de la recherche me permettrait de gagner plus d’argent plus tard<br>-accès à spécialité médicale : je pense que faire de la recherche faciliterait l’accès à une spécialité voulue<br>-statut/prestige : je pense gagner en statut ou en prestige en faisant de la recherche<br>-carrière académique : faire de la recherche me permettrait d’avoir une carrière académique<br>-performance dans la future spécialité : je pense que faire de la recherche me permettrait d’être meilleur dans ma future spécialité<br>-pression extérieure (famille, autre…) : on attend de moi que je fasse de la recherche plus tard<br>-disponibilité/soutien pour future famille : je pense que faire de la recherche me permettrait d’être plus disponible pour ma famille<br>-disponibilité pour autres centres d’intérêts : je pense que faire de la recherche me permettrait d’être plus disponible pour d’autres activités personnelles ou professionnelles<br>-plaisir/intérêt envers la recherche : faire de la recherche me procurerait du plaisir au travail<br>-diversification professionnelle : faire de la recherche me permettrait de diversifier mon travail professionnel<br>-autre : |
| **RA-O9** | 40 | Quelle serait votre raison secondaire à entamer une activité de recherche ? | Idem Q39 |

| | | | | |
|---|---|---|---|---|
| **RA-O10** | 41 | Quelle serait votre raison principale à NE PAS entamer une activité de recherche ? | -argent : je pense que faire de la recherche me ferait gagner moins d'argent plus tard<br>-accès à spécialité médicale : je pense que faire de la recherche compliquerait l'accès à une spécialité voulue<br>-statut/prestige : je pense perdre en statut ou en prestige en faisant de la recherche<br>-pression extérieure (famille, autre…) : on attend de moi que je sois médecin plus tard, pas que je m'engage dans autre chose.<br>-disponibilité/soutien pour future famille : je pense que faire de la recherche m'empêcherait d'être assez disponible pour ma famille<br>-disponibilité pour future spécialité : je pense que faire de la recherche m'empêcherait d'être assez disponible pour ma future spécialité<br>-disponibilité pour autres centres d'intérêts : je pense que faire de la recherche m'empêcherait d'être assez disponible pour d'autres activités personnelles ou professionnelles<br>-plaisir/intérêt envers la recherche : je ne pense pas prendre de plaisir à faire de la recherche plus tard<br>-autre : | |
| **RA-O11** | 42 | Quelle serait la raison secondaire à NE PAS entamer une activité de recherche ? | | Idem Q41 |

**SD : données socio-démographiques**

**SC : choix de spécialité**

**RE : expérience passée**

**R-A : conscience envers la recherche**

**R-V : valeur envers la recherche**

**RA-A : conscience envers une activité de recherche**

**RA-V : valeur envers une activité de recherche**

**RA-E : attente de réussite envers une activité de recherche**

**RA-C : cout envers une activité de recherche**

**RA-I : intention pour une future activité de recherche**

**RA-O : autres questions concernant une future activité de recherche**

**English version, translated from French for the purpose of publishing.**

| ID | ORDER (#) | QUESTIONS | ANSWERS |
|---|---|---|---|
| **SD1** | 1 | How old are you ? | |
| **SD2** | 2 | What is your gender ? | M ; F |
| **SD3** | 3 | To which university are you registered? | UCLouvain ; ULB ; ULg |
| **SD4** | 4 | What is the highest level of education obtained by your mother? | Primary ; Lower secondary ; Upper secondary ; Bachelor's or equivalent ; Master's or equivalent ; Doctor or equivalent ; other: |
| **SD5** | 5 | What is the highest level of education obtained by your father? | Same as #4 |
| **SC1** | 6 | Which specialty did you choose? | |
| **(SC2)** | 7 | Have you hesitated with family medicine? | 1= Not at all ; 2= slightly ; 3= moderately ; 4= strongly ; 5= very strongly |
| **SC3** | 8 | Have you hesitated with another specialty (besides family medicine) ? | 1= Not at all ; 2= slightly ; 3= moderately ; 4= strongly ; 5= very strongly |
| **(SC4)** | 9 | With which specialty (besides family medicine) have you most hesitated ? | |
| **RE1** | 10 | Have you ever considered carrying out a research activity during your medical studies? | 1= Not at all ; 2= slightly ; 3= moderately ; 4= strongly ; 5= very strongly |
| **RE2** | 11 | Did you carry out any research activity during your medical studies? | Yes ; No |

| | | | |
|---|---|---|---|
| **(RE3)** | 12 | What kind of research activity was this? | |
| **(RE4)** | 13 | What is your general appreciation of this research experience ? | 1= Very negative ; 2= negative ; 3= moderate ; 4= positive ; 5= very positive |
| | | **PERCEPTION OF RESEARCH**<br>This section is about the way you perceive research as it is broadly undertaken in your future specialty.<br>By research, we mean a field of activity whose purpose is to increase knowledge in a particular field, using scientific and rigorous methods, in the laboratory or "in the field". | |
| | | Indicate your level of agreement for each element below : | |
| **R-A1** | 14 | I think research is achieved in the field of my future specialty | 1= strongly disagree ; 2= disagree ; 3= slightly disagree ; 4= slightly agree ; 5= agree ; 6= strongly agree |
| **R-V1** | 15 | It is important for me that research is done in the field of my future specialty | Same as #14 |
| **R-V2** | 16 | I show interest towards research within the field of my future specialty | Same as #14 |
| **R-V3** | 17 | I think research performed within the field of my future specialty will be useful for my clinical activity | Same as #14 |
| | | **PERCEPTION OF A RESEARCH ACTIVITY**<br>This section is about the way you perceive a research activity that you would potentially achieve in the future.<br>By research activity, we mean a research project for which you are at the origin of the scientific questioning, and in which you intervene at the different steps of the research process (from the conception of the project to the communication of the results) | |
| | | Indicate your level of agreement for each element below : | |
| **RA-A1** | 18 | I think there are opportunities to conduct research in the field of my future specialty | Same as #14 |
| **RA-E1** | 19 | I feel capable of achieving a research activity | Same as #14 |

| | | | |
|---|---|---|---|
| **RA-E2** | 24 | I believe I could successfully carry out a research activity | Same as #14 |
| **RA-E3** | 27 | I am certain I could understand how to conduct a research activity | Same as #14 |
| **RA-V1** | 22 | Performing a research activity would be important for me | Same as #14 |
| **RA-V2** | 20 | I would be interested in performing a research activity | Same as #14 |
| **RA-V3** | 26 | I think conducting a research activity would be useful for my activity/project/professional goal | Same as #14 |
| **RA-C1** | 21 | Conducting a research activity would require too much time | Same as #14 |
| **RA-C2** | 28 | Given all I will have to do in my personal or professional life, I won't have enough time to carry out a research activity | Same as #14 |
| **RA-C3** | 23 | I would feel incapable of devoting the necessary amount of time to achieve a research activity | Same as #14 |
| **RA-C4** | 25 | I would have to give up too many things if I wanted to achieve a research activity | Same as #14 |
| | | **INTENTION FOR A FUTURE RESEARCH ACTIVITY** | |
| | | Indicate your level of agreement for each element below : | |
| **RA-I1** | 29 | I want to perform a research activity in the future | Same as #14 |
| **RA-I2** | 30 | I intend to perform a research activity in the future | Same as #14 |
| **RA-I3** | 31 | I have planned to achieve a research activity in the future | Same as #14 |
| **RA-O1** | 32 | How do you estimate your chances of carrying out a research activity in the future? | 1= extremely low ; 2= very low ; 3= pretty low ; 4= average ; 5= pretty high ; 6= very high ; 7= extremely high |
| **RA-O2** | 33 | Have you already planned to carry out a research activity in the future? | Yes ; No ; Other |
| **RA-O3** | 34 | When would you prefer to carry out a research activity? | During specialization ; at the beginning of my career ; during my career ; at the end of my career ; other |

| | | | |
|---|---|---|---|
| **RA-O4** | 35 | At what degree of working time would you prefer to carry out a research activity? | <1/10 ; 1/10 ; 2/10 ; 3/10 ; 4/10 ; 5/10 ; 6/10 ; 7/10 ; 8/10 ; 9/10 ; 10/10 ; autre : |
| **RA-O5** | 36 | How would you prefer to carry out a research activity ? | Unique professional activity ; main professional activity with a clinical activity on the side ; secondary professional activity with a main clinical activity ; other |
| **RA-O6** | 37 | In what kind of working environment would you prefer achieving a research activity ? | University ; Professional scientific society ; Private sector (example : pharmaceutical, other) ; Practice, network or hospital-based ; Other : |
| **RA-O7** | 38 | What kind of research activity would you prefer achieve ? | Research-focused scholarship activity ; Doctoral thesis ; Professional activity (unique or complementary) ; Other |
| **RA-O8** | 39 | What would be your main reason for starting a research activity? | - money: I think doing research would make me gain more money later<br>- access to a medical specialty: I think doing research would facilitate access to a desired specialty<br>- status / prestige: I think I will gain status or prestige by doing research<br>- academic career: doing research would allow me to have an academic career<br>- performance in the future specialty: I think doing research would allow me to be better in my future specialty<br>- external pressure (family, other ...): I am expected to do research later<br>- availability / support for future family: I think that doing research would allow me to be more available for my family<br>- availability for other interests: I think doing research would allow me to be more available for other personal or professional activities<br>- pleasure / interest in research: doing research would give me pleasure at work<br>- professional diversification: doing research would allow me to diversify my professional work |

| | | | |
|---|---|---|---|
| | | | - other: |
| **RA-O9** | 40 | What would be your secondary reason for starting a research activity? | Same as #39 |
| **RA-O10** | 41 | What would be your main reason for NOT starting a research activity? | - money: I think doing research would make me gain less money later<br>- access to a medical specialty: I think doing research would complicate access to a desired specialty<br>- status / prestige: I think I will lose status or prestige by doing research<br>- external pressure (family, other ...): I am expected to be a doctor later, not to get involved in something else.<br>- availability / support for future family: I think doing research would prevent me from being sufficiently available for my family<br>- availability for future specialty: I think doing research would prevent me from being sufficiently available for my future specialty<br>- availability for other interests: I think doing research would prevent me from being sufficiently available for other personal or professional activities<br>- pleasure / interest in research: I don't think I will enjoy doing research later<br>- other: |
| **RA-O11** | 42 | What would be your secondary reason for NOT starting a research activity? | Same as #41 |

**SD : socio-demographics**

**SC : specialty choice**

**RE : research experience**

**R-A : research awareness**

**R-V : research value**

**RA-A : research activity awareness**

**RA-V : research activity value**

**RA-E : research activity expectancy**

**RA-C : research activity cost**

**RA-I : research activity intention**

**RA-O : other questions regarding a future research activity**
