## appendix 2 for "Expectancy-Value-Cost motivational theory to explore final year medical students’ research intentions and past research experience: a multicentre cross-sectional questionnaire study"

**APPENDIX 2 – EXPLORATORY FACTOR ANALYSIS RESULTS**

The exploratory factor analysis with oblique rotation (direct oblimin) was conducted on 15 items of the questionnaire. Sample adequacy for the analysis was confirmed with a KMO = 0.88, and with all KMO values of individual items > 0.81. The determinant of the correlation matrix (R-matrix) was over 0.00001, which indicated the absence of a major multicollinearity issue. Three factors had eigenvalues over Kaiser's criterion of 1, but a fourth factor with an eigenvalue of 0.970 was retained based on our interpretation of the scree plot. In combination, the four-factor model explained 75.3% of variance with 6.5% of minimal variance explained by a factor.

**Total Variance Explained**

| | Initial Eigenvalues | | | Extraction Sums of Squared Loadings | | | Rotation Sums of Squared Loadings[a] |
|---|---|---|---|---|---|---|---|
| Factor | Total | % of Variance | Cumulative % | Total | % of Variance | Cumulative % | Total |
| 1 | 7,216 | 48,108 | 48,108 | 6,917 | 46,116 | 46,116 | 5,075 |
| 2 | 2,018 | 13,455 | 61,563 | 1,650 | 11,000 | 57,116 | 4,380 |
| 3 | 1,093 | 7,285 | 68,848 | ,779 | 5,195 | 62,312 | 5,086 |
| 4 | ,970 | 6,467 | 75,315 | ,657 | 4,379 | 66,691 | 2,697 |
| 5 | ,637 | 4,249 | 79,564 | | | | |
| 6 | ,544 | 3,627 | 83,191 | | | | |
| 7 | ,499 | 3,329 | 86,520 | | | | |
| 8 | ,419 | 2,792 | 89,311 | | | | |
| 9 | ,375 | 2,499 | 91,811 | | | | |
| 10 | ,338 | 2,252 | 94,063 | | | | |
| 11 | ,253 | 1,684 | 95,747 | | | | |
| 12 | ,200 | 1,330 | 97,077 | | | | |
| 13 | ,184 | 1,225 | 98,302 | | | | |
| 14 | ,135 | ,900 | 99,202 | | | | |
| 15 | ,120 | ,798 | 100,000 | | | | |

Extraction Method: Principal Axis Factoring.

a. When factors are correlated, sums of squared loadings cannot be added to obtain a total variance.

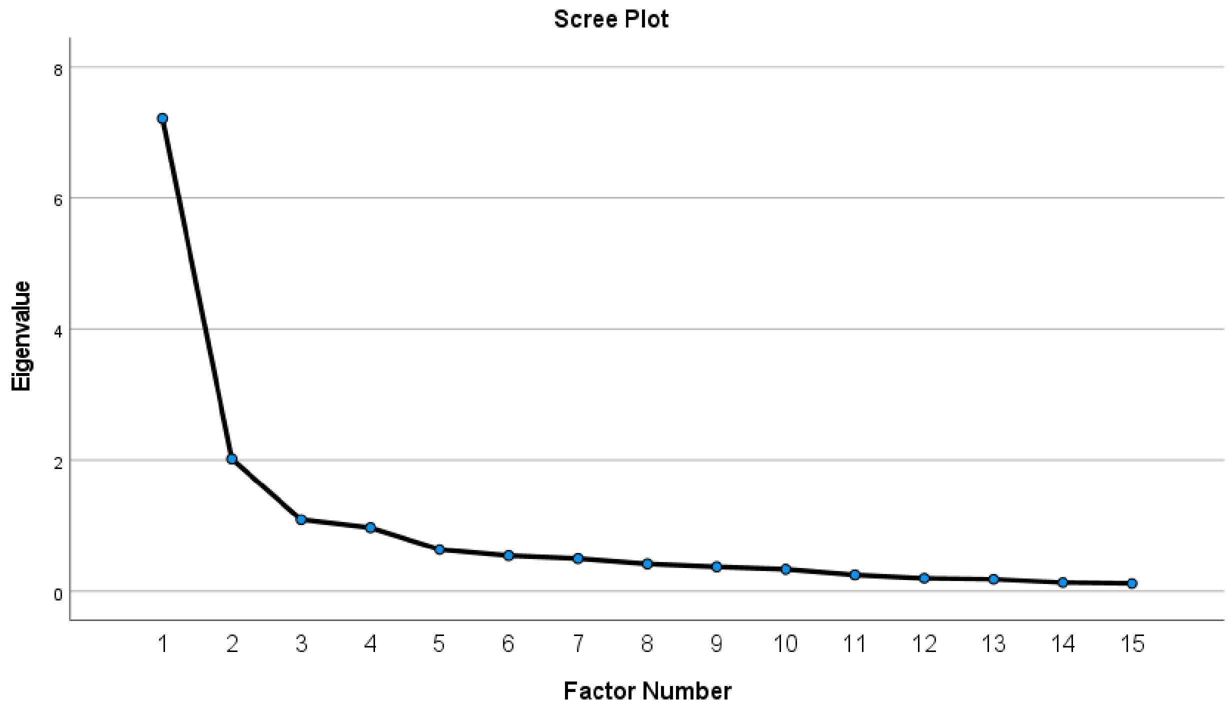


The table below shows the factor correlation matrix. Results confirm correlation between factors and justify the use of oblique rotation.

**Factor Correlation Matrix**

| Factor | 1 | 2 | 3 | 4 |
|---|---|---|---|---|
| 1 | 1,000 | ,352 | ,604 | ,361 |
| 2 | ,352 | 1,000 | ,494 | ,285 |
| 3 | ,604 | ,494 | 1,000 | ,321 |
| 4 | ,361 | ,285 | ,321 | 1,000 |

Extraction Method: Principal Axis Factoring.

Rotation Method: Oblimin with Kaiser Normalization.
