## appendix 3 for "Expectancy-Value-Cost motivational theory to explore final year medical students’ research intentions and past research experience: a multicentre cross-sectional questionnaire study"

### APPENDIX 3 - SPEARMAN'S RANK-ORDER CORRELATIONS RESULTS

*Table - Spearman's rank-order correlation results*

| | Consid [N] | Apprec [Y] | A | E | V | C |
|---|---|---|---|---|---|---|
| **Considered doing research** [N] | 1.000 | | | | | |
| **Research experience appreciation** [Y] | | 1.000 | | | | |
| **Awareness (A)** | 0.346** | 0.581** | 1.000 | | | |
| **Expectancy for success (E)** | 0.430** | 0.626** | 0.522** | 1.000 | | |
| **Value (V)** | 0.640** | 0.633** | 0.565** | 0.661** | 1.000 | |
| **Cost (C)** | -0.445** | -0.422** | -0.368** | -0.581** | -0.611** | 1.000 |
| **Intention (I)** | 0.695** | 0.573** | 0.545** | 0.670** | 0.894** | -0.624** |

[N] n = 125 (students without research experience) [Y] n = 70 (students with research experience)

** correlation is significant at 0.01 level (two-tailed)
