## appendix 4 for "Expectancy-Value-Cost motivational theory to explore final year medical students’ research intentions and past research experience: a multicentre cross-sectional questionnaire study"

### APPENDIX 4 – HIERARCHICAL MULTIPLE REGRESSION ANALYSIS

Assumptions for multiple regression analysis were verified. The design of the study implies independence of observations (no autocorrelation). Scatterplots showed a linear relationship between the dependent and independent variables. Residuals were normally distributed confirming multivariate normality. No multicollinearity problem was identified as VIF values were below 3. There was homoscedasticity, as assessed by visual inspection of a plot of studentized residuals versus unstandardized predicted values.

We detected one possible significant outlier with a standardized residual of -3.34. Cook's distance for this case was 0.147 (highest of dataset). By comparing regression coefficients with and without this case, we saw impact on Cost and Expectancy results. Nevertheless, in our opinion, these differences were not sufficient enough to exclude this case.

The addition of research experience during medical school (Model 3) to a model including profile variables (age, gender, university – model 1) and medical specialty (model 2) led to a statistically significant increase in explained variance of 10.4% ($\Delta R^2$). The addition of motivational factors (Models 4, 5, 6, 7) led, altogether, to a statistically significant increase in explained variance of 57.5%.

| | Model 1 B | Model 1 Beta | Model 2 B | Model 2 Beta | Model 3 B | Model 3 Beta | Model 4 B | Model 4 Beta | Model 5 B | Model 5 Beta | Model 6 B | Model 6 Beta | Model 7 B | Model 7 Beta |
|---|---|---|---|---|---|---|---|---|---|---|---|---|---|---|
| (Intercept) | 1,99 | | 1,69 | | 0,74 | | -2,30 | | 0,73 | | 0,67 | | -0,71 | |
| Age (years) | 0,04 | 0,05 | 0,07 | 0,09 | **0,10*** | **0,13** | 0,08 | 0,10 | **0,09*** | **0,11** | 0,05 | 0,06 | 0,04 | 0,05 |
| Gender | | | | | | | | | | | | | | |
| Female | | | | | | | | | | | | | | |
| Male | 0,50 | 0,17 | 0,34 | 0,11 | 0,28 | 0,09 | 0,27 | 0,09 | 0,24 | 0,08 | 0,00 | 0,00 | 0,08 | 0,03 |
| University | | | | | | | | | | | | | | |
| ULB | | | | | | | | | | | | | | |
| UCLouvain | -0,12 | -0,04 | 0,02 | 0,01 | -0,06 | -0,02 | -0,09 | -0,03 | -0,07 | -0,02 | -0,09 | -0,03 | -0,11 | -0,04 |
| ULg | 0,15 | 0,04 | 0,26 | 0,06 | 0,15 | 0,03 | 0,17 | 0,04 | 0,13 | 0,03 | 0,16 | 0,04 | 0,10 | 0,02 |
| Speciality | | | | | | | | | | | | | | |
| Hospital-based | | | | | | | | | | | | | | |
| Family Medicine | | | **-1,02**** | **-0,34** | **-0,86**** | **-0,29** | **-0,40*** | **-0,13** | -0,30 | -0,10 | -0,25 | -0,08 | -0,08 | -0,03 |
| RE during medical school | | | | | | | | | | | | | | |
| No | | | | | | | | | | | | | | |
| Yes | | | | | **0,98**** | **0,33** | **0,87**** | **0,30** | **0,53*** | **0,18** | **0,44*** | **0,15** | 0,16 | 0,05 |
| Awareness | | | | | | | **0,68**** | **0,43** | **0,46**** | **0,29** | **0,30*** | **0,19** | 0,02 | 0,02 |
| Cost | | | | | | | | | **-0,57**** | **-0,45** | **-0,40**** | **-0,31** | **-0,14*** | **-0,11** |
| Expectancy | | | | | | | | | | | **0,40**** | **0,34** | **0,12*** | **0,10** |
| Value | | | | | | | | | | | | | **0,77**** | **0,72** |
| $R^2$ | 0.037 | | **0.149** | | **0.253** | | **0.412** | | **0.568** | | **0.624** | | **0.828** | |
| F | 1.85 | | **6.60**** | | **10.62**** | | **18.78**** | | **30.52**** | | **34.09**** | | **88.88**** | |
| $\Delta R^2$ | / | | **0.111** | | **0.104** | | **0.159** | | **0.155** | | **0.056** | | **0.205** | |

* $p < 0.05$ ; ** $p < 0.001$

The order of introduction of the independent variables was established based on the results of the Spearman's rank-order correlations results (from lowest to highest).
